# Real-World Dose Modifications for FOLFIRINOX in Pancreatic Cancer: Evaluating the Feasibility of A Machine-Learning Framework

**DOI:** 10.64898/2026.04.27.26350002

**Authors:** Akanksha Dua, Ziad Obermeyer, Atul Butte, Travis Zack

## Abstract

**Background:** FOLFIRINOX is a cornerstone regimen for eligible patients with pancreatic ductal adenocarcinoma (PDAC), but its clinical benefit is limited by substantial toxicity and frequent dose modification. In real-world practice, dose modifications are often individualized, and the clinical factors associated with these decisions remain incompletely characterized.

**Objective:** To develop and evaluate an electronic medical record (EMR)–based machine-learning framework for modeling cycle-specific FOLFIRINOX dose modification decisions in patients with PDAC.

**Methods:** We included patients with PDAC who received FOLFIRINOX at UCSF oncology clinics between November 2011 and December 2023. Predictors included demographic, clinical, laboratory, and treatment variables derived from the EMR. Logistic regression, random forest, and XGBoost models were trained using group-based 5-fold cross-validation to predict cycle-specific dose modifications for 5-fluorouracil, irinotecan, and oxaliplatin. Model performance was evaluated using area under the receiver operating characteristic curve.

**Results:** The cohort included 514 patients receiving FOLFIRINOX across 5,041 treatment cycles. The mean age was 59 years, 60% of patients were White, 41% had a history of smoking, and patients received a median of 6 chemotherapy cycles. More than 60% of patients required at least one dose modification during treatment. XGBoost demonstrated the highest performance across component drugs, with AUCs ranging from 0.53 to 0.70. Clinically plausible predictors of irinotecan and oxaliplatin dose modification included hepatic and renal function markers, cumulative drug exposure, treatment-related symptoms, and demographic or behavioral characteristics.

**Conclusion:** We developed an EMR-based machine-learning framework to model real-world FOLFIRINOX dose modification and identified clinically plausible, routinely available predictors, particularly for irinotecan and oxaliplatin. Variable model performance suggests that dosing decisions are only partially captured by structured EMR data, highlighting both the limitations of current data-driven approaches and clinical domains where ML-based models may support individualized dosing and toxicity surveillance. Future informatics efforts should incorporate dose-modification rationale, patient-reported and functional outcomes, and validation across diverse practice settings.

## Background

Pancreatic ductal adenocarcinoma (PDAC) is an aggressive malignancy with a 5-year survival rate of approximately 10% and is projected to become the second leading cause of cancer-related mortality by 2030.^1,2^ Most patients present with advanced disease, limiting eligibility for surgical resection, and systemic chemotherapy remains the mainstay of treatment. However, in real-world practice, treatment is frequently de-escalated or discontinued due to toxicity and limited tolerability.^3,4,5^ Despite this, there is no standardized framework to guide therapy modification, resulting in substantial variation between planned and delivered dosing and idiosyncratic treatment patterns.^6^

FOLFIRINOX, a combination regimen of 5-fluorouracil, irinotecan, and oxaliplatin, is a cornerstone therapy for patients with locally advanced or metastatic PDAC and is associated with improved survival.^7,8^ However, its use is limited by a high toxicity burden, necessitating frequent dose modifications even among patients with good performance status.^9,10^ While several studies have evaluated different FOLFIRINOX dosing strategies, no consensus exists on how to best balance toxicity-related concerns while retaining clinical benefit.^10,11^ In practice, real-world dosing decisions vary widely and may be shaped by patient preference, institution guidelines, provider-specific risk assessment, as well as other implicit factors. Known disparities in the clinical burden and management of PDAC by race and access to care further suggest that unmeasured social and clinical determinants influence dose modification patterns and may be challenging to capture systematically.^12–15^ Therefore, it is important to leverage existing data repositories to better elucidate the clinical and demographic drivers of sub-optimal dose-effect relationships.

Given the aggressiveness of the disease, and toxicity of regimens, treatment options after progression is not an option for many patients, and overall survival (OS) after treatment discontinuation is only 1-3 months.^16–18^ This makes it imperative to select the appropriate dosing regimen in the first line. Identifying predictors of dose modification may enable earlier intervention, reduce high-grade toxicities, and improve treatment continuity. However, dose modification decisions remain largely empirical, with limited understanding of which patients require adjustments and when these should occur. This variability also limits the ability to evaluate outcomes across diverse patient populations.^19^

Given these inconsistencies in real world treatment patterns and the absence of universal dose modification protocols, we can leverage data-driven machine-learning (ML) models to characterize treatment landscape more comprehensively, uncover patterns in decision-making, and evaluate whether predictive frameworks can guide optimal dosing strategies. These approaches offer a potential framework to capture complex, non-linear relationships between patient characteristics, treatment exposures, and clinical outcomes. While several studies have applied ML approaches to predict disease risk and treatment outcomes in PDAC, few have focused on modeling real-world chemotherapy dosing decisions.^20–22^

This study represents a first step toward addressing gaps in real-world FOLFIRINOX dosing by developing an ML framework to model dose modification decisions using longitudinal data from a tertiary academic center. We evaluate the feasibility of ML-based approaches to characterize real-world dosing decisions and identify clinically relevant predictors that may inform more individualized, data-driven treatment strategies. More broadly, this work explores how real-world data can inform patient-centric treatment approaches that balance efficacy, safety, and quality of life, with implications for clinical decision-making, pharmacologic research, and regulatory evaluation.

## Methods

### Data source

The current study was a retrospective, longitudinal analysis conducted using de-identified EPIC-based electronic medical record (EMR) data from University of California – San Francisco (UCSF). Data include the breadth of the EMR, including, but not limited to structured elements such as International Classification of Disease (ICD) codes, National Drug Codes (NDC) codes, Healthcare Procedure Coding System (HCPCS) codes, clinical laboratory test results, in addition to unstructured information extracted from medical charts, notes, problem lists, and encounters. Treatment information, including cycle, dose, and scheduling information were collected. Patient covariates such as demographics, comorbidities, and laboratory data were extracted. This has enabled the generation of a comprehensive dataset that includes potential causal variables, confounders, and outcomes in the treatment of PDAC.

Given that the current study involves secondary analysis of de-identified data, it was determined to be exempt from Institutional Review Board (IRB) review under Exemption Category 4.

### Study population

The study population consisted of patients that attended gastrointestinal (GI) medical oncology clinics at UCSF between 2011 and 2023, with at least one confirmed diagnosis of PDAC using ICD-CM codes (ICD 9-CM: 157.x, ICD-10-CM: C25.x), a visit to our GI medical oncology department, and a mention of PDAC in the clinical notes from this department. Patients were further required to have at least one documented administration of FOLFIRINOX, which includes infusion with 5-fluorouracil, irinotecan, and oxaliplatin. No other demographic or disease-specific exclusion criteria were applied for the study.

### Study design

The study period spanned from November 2011 – December 2023. The index date for the study was defined as the date of the first documented administration of FOLFIRINOX (i.e., first cycle date) following a diagnosis of PDAC. The observation period ranged from the cycle date until the end of treatment regimen, end of data availability, or death. Baseline characteristics included variables that were defined at the start of the study, on or before the first cycle date.

### Demographic, clinical, and laboratory predictors

Patient demographic data were collected on index date and included age, race, ethnicity, marital status, and smoking history. Diagnosis data were collected prior to cycle date of interest (baseline diagnoses). The most frequently occurring diagnoses were extracted (i.e., >25% of the patient population) and included as clinical predictors. Laboratory measures that were present in >80% of patients were summarized and included as predictors. All laboratory measures that occurred before the corresponding cycle date were included as predictors for that cycle to more adequately capture any fluctuations in patients’ clinical trajectories. A list of all predictors is included as **Supplemental Table 1**.

### Definition of outcomes

Dose modifications (i.e., reductions) were defined as any instance where there was a decrease (of at least 10%) in the dose of the drug compared to the previous cycle or instances where the dose of the drug was reduced to zero (drug discontinuation) as compared to the previous cycle.

### Statistical analysis

#### Baseline characteristics

Demographic and clinical characteristics were summarized for the overall population, and for subgroups of patients receiving zero or 1+ dose modifications or reductions. Continuous variables were described using means and standard deviations (SDs) and categorical variables were described using frequencies and percentages. Cohorts were compared using chi-square tests for categorical variables, and Wilcoxon rank-sum tests for continuous variables.

### Dosage patterns

Distribution of dosing patterns were descriptively summarized for the overall population by cycle of treatment and treatment received (i.e., 5-fluorouracil, irinotecan, and oxaliplatin). Data across all treatment cycles were summarized to evaluate the proportion of patients receiving full doses, or any dose modifications and discontinuations. Data were then normalized to sum up to 100%. Results for the first 30 cycles were visualized via stacked bar graphs.

### Machine-learning framework

A machine-learning framework was generated for predicting the outcomes of interest using a group-based 5-fold cross validation for logistic regression, random forest, and Extreme Gradient Boosting (XGB) models. The specific model parameters are noted in **Supplemental Table 2**. Synthetic Minority Oversampling Technique (SMOTE) regularization was applied to address class imbalance issues and improve accuracy of estimates due to low overall occurrence of outcomes.^23^ Receiver Operating Characteristic Area Under the Curve (ROC-AUC) curves were plotted to evaluate and summarize model performance across all classification thresholds. Mean estimates and associated 95% confidence intervals (CIs) for feature importance were obtained from bootstrap sampling (n =100 iterations). The Bonferroni correction for multiple comparisons was used to generate adjusted p-values to reduce the probability of Type I (false positive) errors.^24^ Shapley Additive Explanations (SHAP) values were calculated to determine the contribution of each feature on model prediction. Features were ranked by their average absolute SHAP values and plotted against bootstrapped model coefficients to evaluate individual as well as overall contributions of variables on the model outcome.

All analyses were conducted using R and Python using a secure server hosted on UCSF. The machine learning models were developed using Sklearn package in Python.

## Results

### Baseline characteristics and dosing patterns

A total of 514 patients with PDAC with at least one confirmed administration of FOLFIRINOX were included in our study. In our study sample, the average age was 58.8 years, 59.5% of patients were White, 20.2% were Asian, 8.0% were of Hispanic or Latino origin (**Table 1**). 41.1% of patients had some history of smoking. Patients received a median of 6.0 chemotherapy cycles (first and third quantiles), with each cycle lasting approximately 2 weeks. Across the three treatment regimens evaluated, 5-fluorouracil had the lowest proportion of dose modifications (26.9%), followed by oxaliplatin (36.0%), and irinotecan (37.0%). 5-fluorouracil also had the lowest proportion of discontinuations (3.5%), followed by irinotecan (11.3%), and oxaliplatin (23.4%). The most commonly reported clinical features at baseline included dehydration (80.2%), followed by nausea (71.0%), liver or bile-duct neoplasm (41.4%), anemia (39.5%), diarrhea (39.1%), and abdominal pain (30.9%). Overall, the mean value for liver function tests were abnormal, including higher levels of alanine transaminase (ALT), aspartate transaminase (AST), alkaline phosphatase (ALP), and bilirubin. Of the patients that had available baseline liver function tests, 38.8% had an ALT >35, 32.2% had an AST >35, 43.5% had an ALP >100 and 24.5% had a total bilirubin >1.0. Of the patients that had a baseline measure of cancer antigen 19-9, 76.0% had an abnormally elevated value of >37 U/L, with a median CA 19-9 value of 491. Dosing patterns for the three medications (5-fluorouracil, irinotecan, oxaliplatin) are summarized in **Figure 1**. Modifications and discontinuations most commonly occurred in cycle 12 (5-fluorouracil: 12%, irinotecan : 20%; oxaliplatin: 33%). For oxaliplatin, dose discontinuations were more frequent compared to other treatment regimens, ranging from 4% to 18%, whereas dose discontinuations ranged from 1-7% for irinotecan, and 1-5% for 5-fluorouracil.

**Figure 1.**
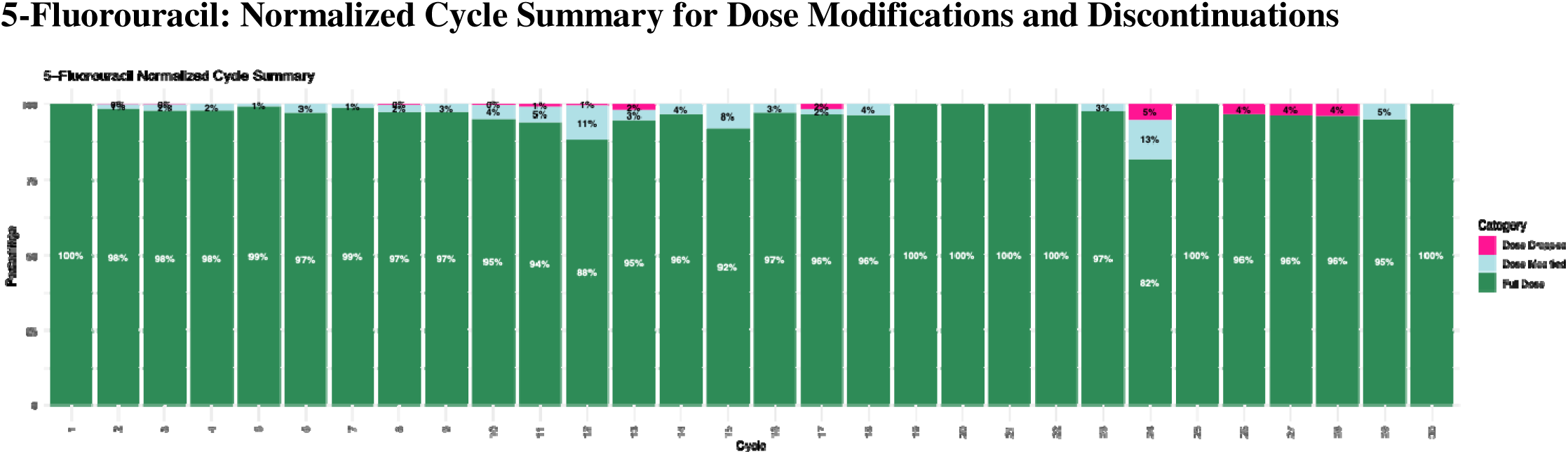
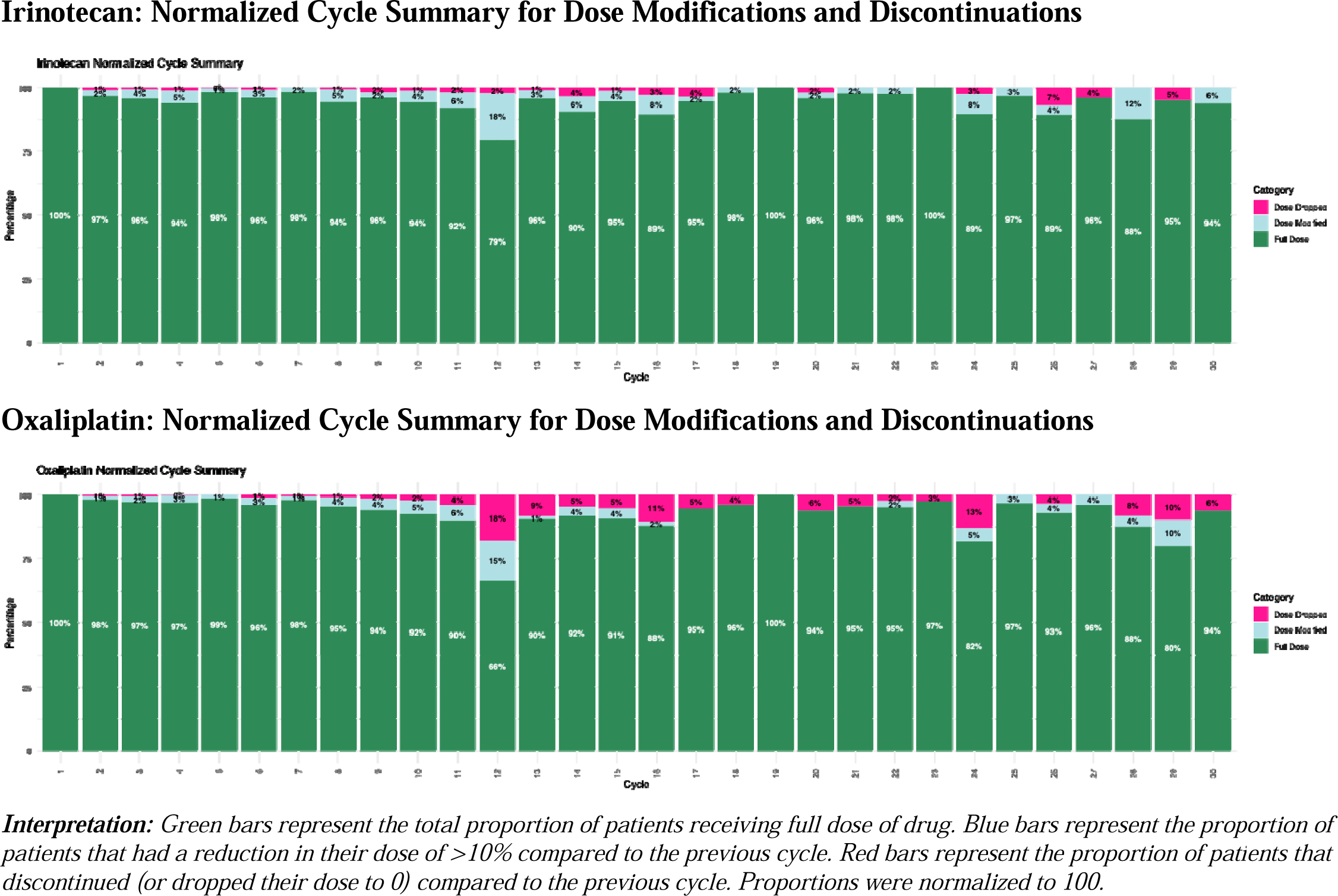
FOLFIRINOX Dose Modifications and Discontinuations.

**Table 1.**
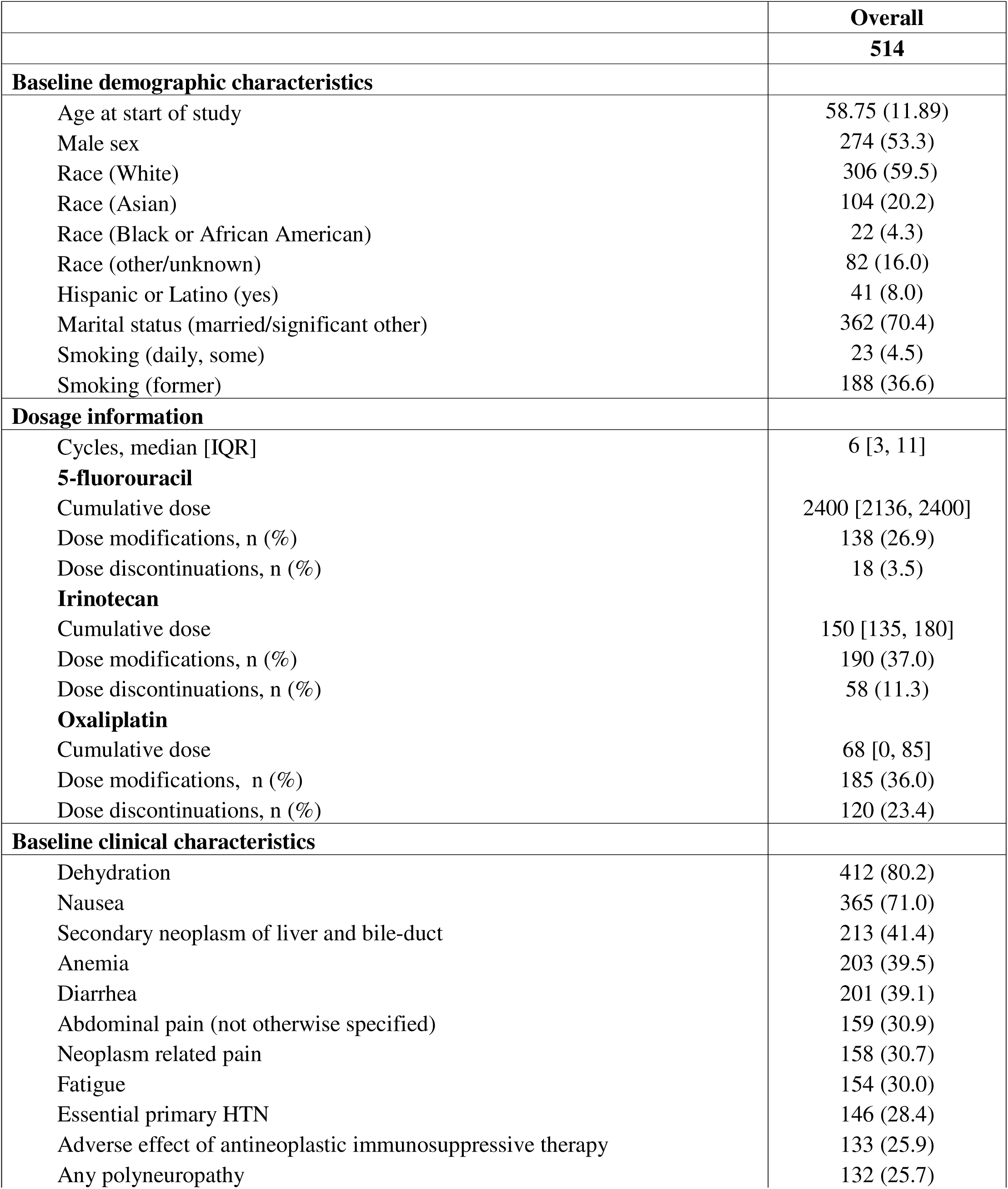
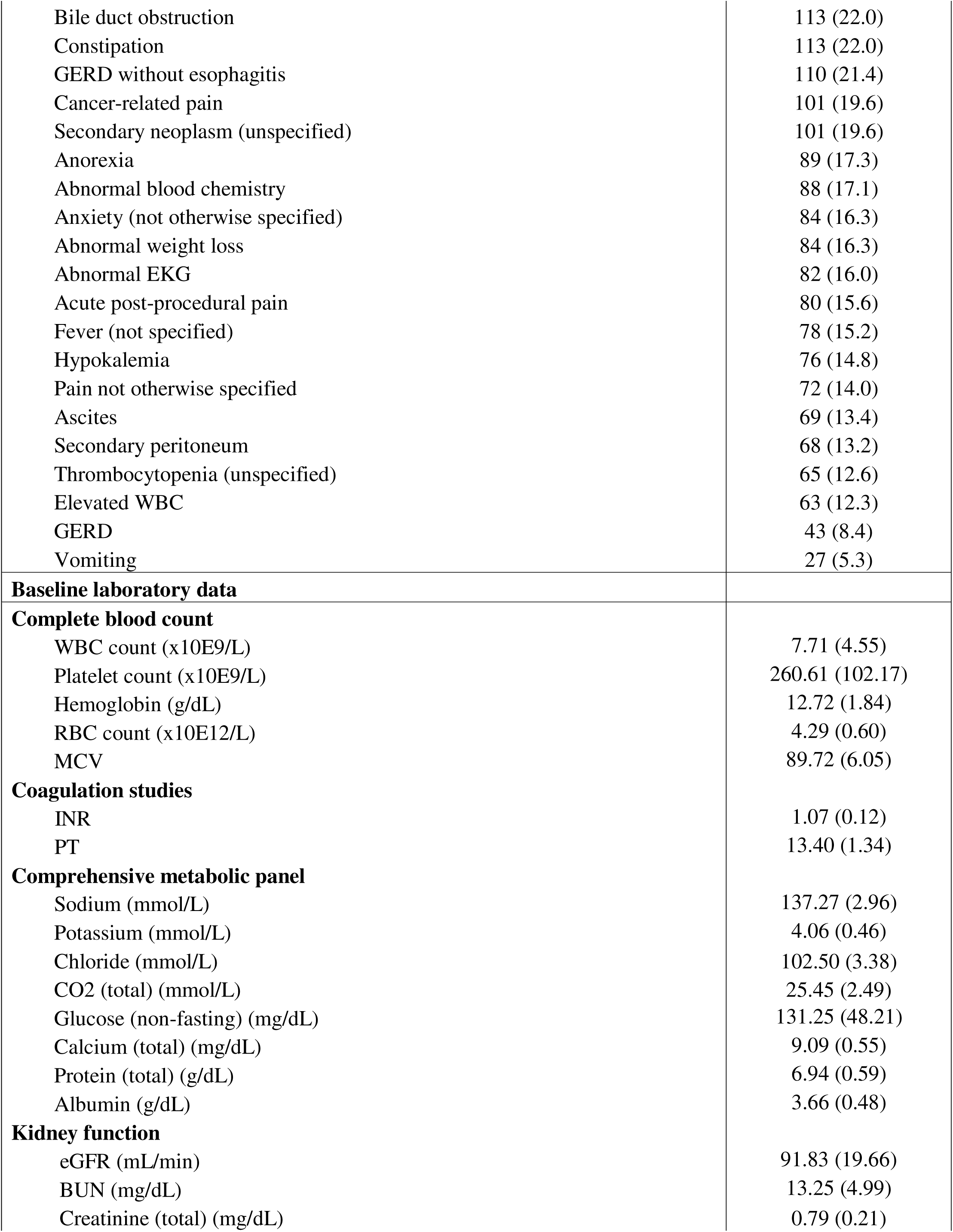
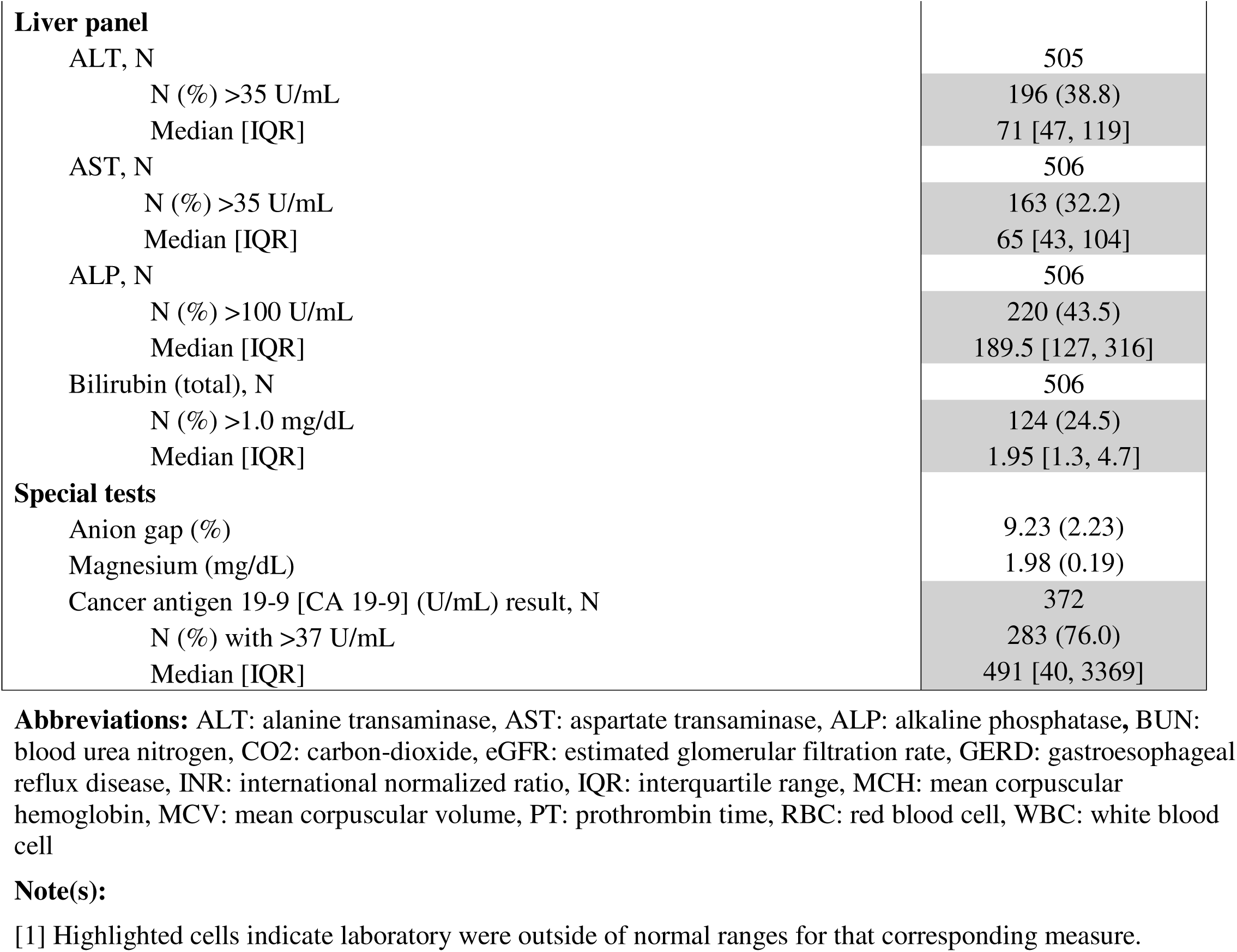
Baseline Characteristics of Patients with PDAC that Attended UCSF Oncology Clinics between 2011 – 2023.

|  | **Overall** |
| --- | --- |
|  | **514** |
| **Baseline demographic characteristics** |  |
| Age at start of study | 58.75 (11.89) |
| Male sex | 274 (53.3) |
| Race (White) | 306 (59.5) |
| Race (Asian) | 104 (20.2) |
| Race (Black or African American) | 22 (4.3) |
| Race (other/unknown) | 82 (16.0) |
| Hispanic or Latino (yes) | 41 (8.0) |
| Marital status (married/significant other) | 362 (70.4) |
| Smoking (daily, some) | 23 (4.5) |
| Smoking (former) | 188 (36.6) |
| **Dosage information** |  |
| Cycles, median [IQR] | 6 [3, 11] |
| **5-fluorouracil** |  |
| Cumulative dose | 2400 [2136, 2400] |
| Dose modifications, n (%) | 138 (26.9) |
| Dose discontinuations, n (%) | 18 (3.5) |
| **Irinotecan** |  |
| Cumulative dose | 150 [135, 180] |
| Dose modifications, n (%) | 190 (37.0) |
| Dose discontinuations, n (%) | 58 (11.3) |
| **Oxaliplatin** |  |
| Cumulative dose | 68 [0, 85] |
| Dose modifications, n (%) | 185 (36.0) |
| Dose discontinuations, n (%) | 120 (23.4) |
| **Baseline clinical characteristics** |  |
| Dehydration | 412 (80.2) |
| Nausea | 365 (71.0) |
| Secondary neoplasm of liver and bile-duct | 213 (41.4) |
| Anemia | 203 (39.5) |
| Diarrhea | 201 (39.1) |
| Abdominal pain (not otherwise specified) | 159 (30.9) |
| Neoplasm related pain | 158 (30.7) |
| Fatigue | 154 (30.0) |
| Essential primary HTN | 146 (28.4) |
| Adverse effect of antineoplastic immunosuppressive therapy | 133 (25.9) |
| Any polyneuropathy | 132 (25.7) |
| Bile duct obstruction | 113 (22.0) |
| Constipation | 113 (22.0) |
| GERD without esophagitis | 110 (21.4) |
| Cancer-related pain | 101 (19.6) |
| Secondary neoplasm (unspecified) | 101 (19.6) |
| Anorexia | 89 (17.3) |
| Abnormal blood chemistry | 88 (17.1) |
| Anxiety (not otherwise specified) | 84 (16.3) |
| Abnormal weight loss | 84 (16.3) |
| Abnormal EKG | 82 (16.0) |
| Acute post-procedural pain | 80 (15.6) |
| Fever (not specified) | 78 (15.2) |
| Hypokalemia | 76 (14.8) |
| Pain not otherwise specified | 72 (14.0) |
| Ascites | 69 (13.4) |
| Secondary peritoneum | 68 (13.2) |
| Thrombocytopenia (unspecified) | 65 (12.6) |
| Elevated WBC | 63 (12.3) |
| GERD | 43 (8.4) |
| Vomiting | 27 (5.3) |
| **Baseline laboratory data** |  |
| **Complete blood count** |  |
| WBC count (x10E9/L) | 7.71 (4.55) |
| Platelet count (x10E9/L) | 260.61 (102.17) |
| Hemoglobin (g/dL) | 12.72 (1.84) |
| RBC count (x10E12/L) | 4.29 (0.60) |
| MCV | 89.72 (6.05) |
| **Coagulation studies** |  |
| INR | 1.07 (0.12) |
| PT | 13.40 (1.34) |
| **Comprehensive metabolic panel** |  |
| Sodium (mmol/L) | 137.27 (2.96) |
| Potassium (mmol/L) | 4.06 (0.46) |
| Chloride (mmol/L) | 102.50 (3.38) |
| CO2 (total) (mmol/L) | 25.45 (2.49) |
| Glucose (non-fasting) (mg/dL) | 131.25 (48.21) |
| Calcium (total) (mg/dL) | 9.09 (0.55) |
| Protein (total) (g/dL) | 6.94 (0.59) |
| Albumin (g/dL) | 3.66 (0.48) |
| **Kidney function** |  |
| eGFR (mL/min) | 91.83 (19.66) |
| BUN (mg/dL) | 13.25 (4.99) |
| Creatinine (total) (mg/dL) | 0.79 (0.21) |

|  |  |  |
| --- | --- | --- |
| **Liver panel** |  |  |
| ALT, N |  | 505 |
| N (%) >35 U/mL |  | 196 (38.8) |
| Median [IQR] |  | 71 [47, 119] |
| AST, N |  | 506 |
| N (%) >35 U/mL |  | 163 (32.2) |
| Median [IQR] |  | 65 [43, 104] |
| ALP, N |  | 506 |
| N (%) >100 U/mL |  | 220 (43.5) |
| Median [IQR] |  | 189.5 [127, 316] |
| Bilirubin (total), N |  | 506 |
| N (%) >1.0 mg/dL |  | 124 (24.5) |
| Median [IQR] |  | 1.95 [1.3, 4.7] |
| **Special tests** |  |  |
| Anion gap (%) |  | 9.23 (2.23) |
| Magnesium (mg/dL) |  | 1.98 (0.19) |
| Cancer antigen 19-9 [CA 19-9] (U/mL) result, N |  | 372 |
| N (%) with >37 U/mL |  | 283 (76.0) |
| Median [IQR] |  | 491 [40, 3369] |

**Abbreviations:** ALT: alanine transaminase, AST: aspartate transaminase, ALP: alkaline phosphatase, BUN: blood urea nitrogen, CO2: carbon-dioxide, eGFR: estimated glomerular filtration rate, GERD: gastroesophageal reflux disease, INR: international normalized ratio, IQR: interquartile range, MCH: mean corpuscular hemoglobin, MCV: mean corpuscular volume, PT: prothrombin time, RBC: red blood cell, WBC: white blood cell

### Descriptive characteristics of patients receiving dose modifications

Patients were classified into two cohorts depending on whether they received zero or 1+ dose modifications and/or discontinuations (**Supplemental Table 3**) during treatment. Of the 514 patients, 186 (36.2%) had zero dose modifications/discontinuations, and 328 (63.8%) received 1+ dose modifications/discontinuations. Compared to patients that had zero dose modifications/discontinuations, patients that received 1+ dose modifications/discontinuations were on average younger (58.2 [12.4] vs. 59.7 [10.9], p = 0.173), less likely to be male (51.8% vs. 55.9%, p = 0.424), and received higher cumulative 5-fluorouracil (2450.46 (397.60) vs. 2212.37 (415.03), p<0.001) and higher cumulative irinotecan doses (151.09 (43.35) vs. 145.91 (44.74), p = 0.199). In terms of baseline clinical characteristics, patients with 1+ dose modifications/reductions were significantly more likely to have had diarrhea (45.7% vs. 27.4%), polyneuropathy (34.8% vs. 9.7%), and fatigue (36.3 vs. 18.8%) (all p<0.001). As expected, typical adverse effects of treatment were more likely in the group who underwent multiple treatment modifications (33.8% vs. 11.8%, p<0.001). Compared to patients that had zero dose modifications/discontinuations, patients that had 1+ dose modification/discontinuation were also more likely to experience adverse outcomes during the study period (i.e., after baseline), including higher rates of dehydration (36.6% vs. 0.0%), polyneuropathy (32.0% vs. 7.0%), and diarrhea (39.6% vs. 21.5%) (all p<0.001). There were fewer deaths in the cohort that had 1+ treatment modification/discontinuation (57.3% vs. 61.8%, p =0.365), and patients were younger at time of death (60.3 vs. 61.4 years, p=0.451), though these were not significant.

### Summary of machine-learning models

Of all the models tested, the XGB classifier was the best model for predicting dose modifications/reductions for 5-fluorouracil (AUC: 0.53), irinotecan (AUC: 0.59) and oxaliplatin (AUC: 0.70 **Table 2**). Results for the best performing model are summarized, unless otherwise specified. If mean AUCs were comparable between random forest and XGB models, then results of XGB classifier models are included for consistency. The graphs representing the AUCs for all of the models are reported in **Figure 2**. Given the poor performance of the 5-fluorouracil model, we did not believe evaluation of features that predict reductions were informative for the current analyses, and have not presented the results here.

**Figure 2.**
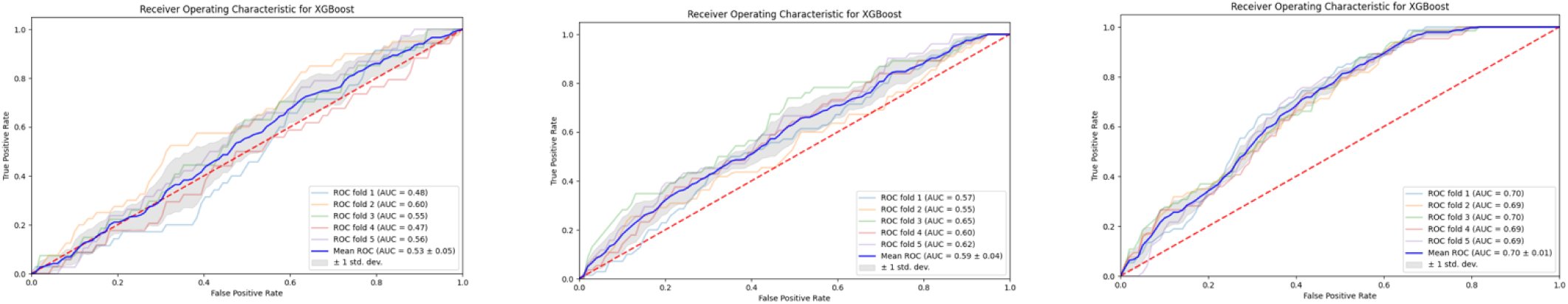
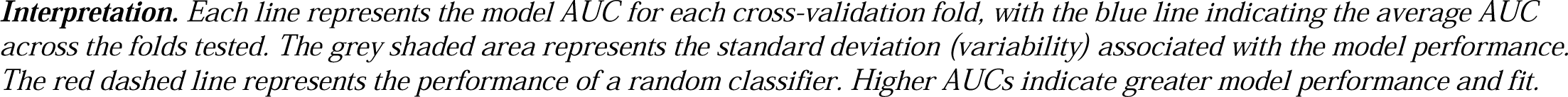
FOLFIRINOX Dose Modification/Reduction – ROC AUC Curves for 5-Fluorouracil, Irinotecan, and Oxaliplatin.

**Table 2.** Summary of FOLFIRINOX Model Outputs Across Outcomes.

| Outcome \ Model | Logistic Regression | Random Forest | XGBoost |
| --- | --- | --- | --- |
| 5-Fluorouracil Dose Modification | 0.52 ± 0.01 | 0.53 ± 0.03 | 0.53 ± 0.05 |
| Irinotecan Dose Modification | 0.55 ± 0.02 | 0.58 ± 0.04 | 0.59 ± 0.04 |
| Oxaliplatin Dose Modification | 0.65 ± 0.02 | 0.68 ± 0.01 | 0.70 ± 0.01 |

### Irinotecan

Features that were significantly associated with irinotecan dose modifications included demographic features such as male sex, smoking status, measures of liver and kidney function such as latest total bilirubin, INR, eGFR, and potassium, baseline magnesium, CO2, and acute postprocedural pain (all adjusted p<0.05). Dosing patterns such as fluorouracil infusion and irinotecan administration were also identified as important features in predicting dose modifications (p<0.05). Male sex, irinotecan administration, smoking status, and total bilirubin were also associated with the highest SHAP values, indicating importance in predicting both individual patients and clusters, as well as in the overall model. Clinical features such as any polyneuropathy, fever, and diarrhea were associated with high SHAP values, but lower feature importance values, indicating that these measures are likely important for certain individuals or subgroups (i.e., individual observations) but not for the overall population (i.e., global model accuracy). Variables with the highest SHAP values and variables with the highest bootstrapped feature importance coefficients (with adjusted p<0.05) are summarized in **Figure 3A**.

**Figure 3A.**
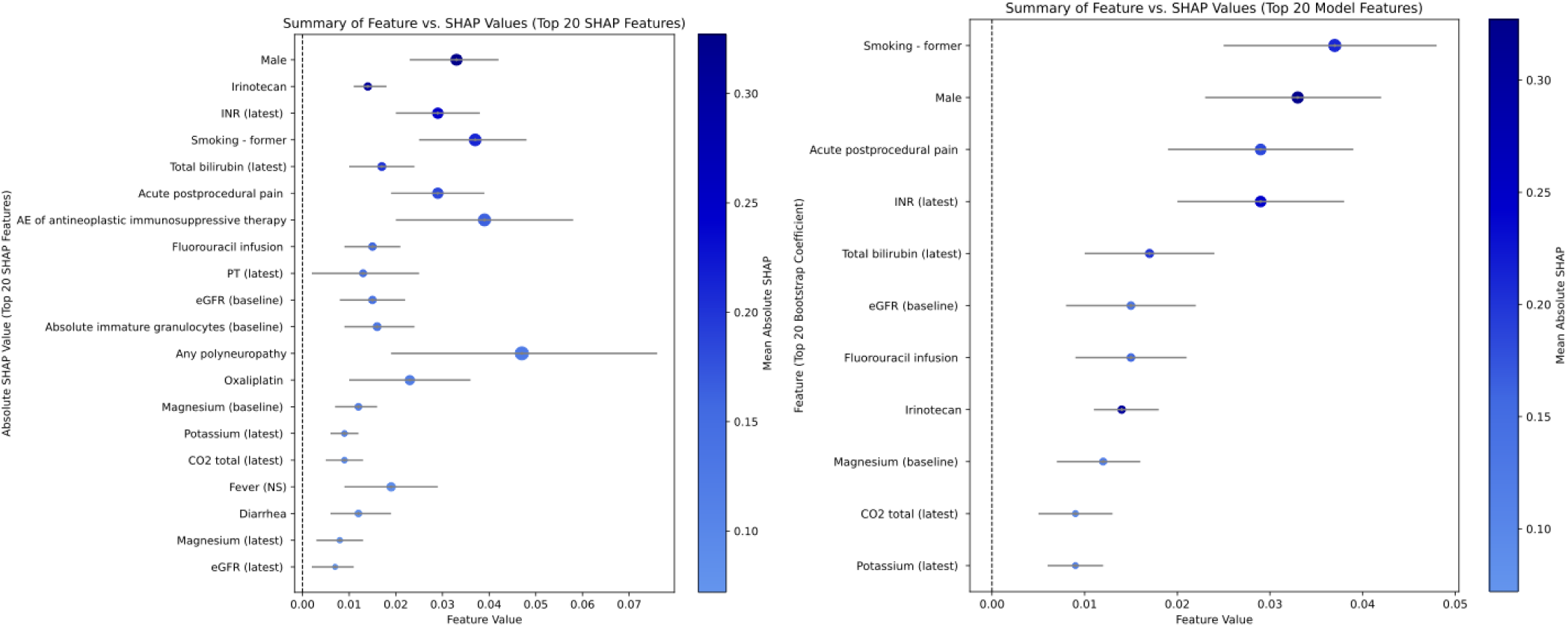
*FOLFIRINOX* Dose Modification/Reduction Feature Importance vs. SHAP Importance for Irinotecan.

### Oxaliplatin

Features that were significantly associated with oxaliplatin dose modification included oxaliplatin and cumulative oxaliplatin doses, clinical features such as essential primary hypertension and diarrhea, demographic features such as smoking status and male sex, and laboratory measures such as baseline potassium, absolute eosinophils, and platelet count (all Bonferroni p<0.05). Oxaliplatin, smoking history, male sex, and essential primary hypertension also had the highest SHAP values (in the top 20%), indicating importance in predicting both individual patients and clusters, as well as in the overall model. Latest PT, INR, and liver function tests (e.g., bilirubin), clinical features such as abdominal pain, fatigue, and secondary peritoneum were also associated with SHAP values in the top 20%, indicating importance in prediction of individual patients or clusters, but lower feature values (none of the associations were significant) indicating lower overall importance in the model. Variables with the highest SHAP values and variables with the highest bootstrapped feature importance coefficients (with adjusted p<0.05) are summarized in **Figure 3B**.

**Figure 3B.**
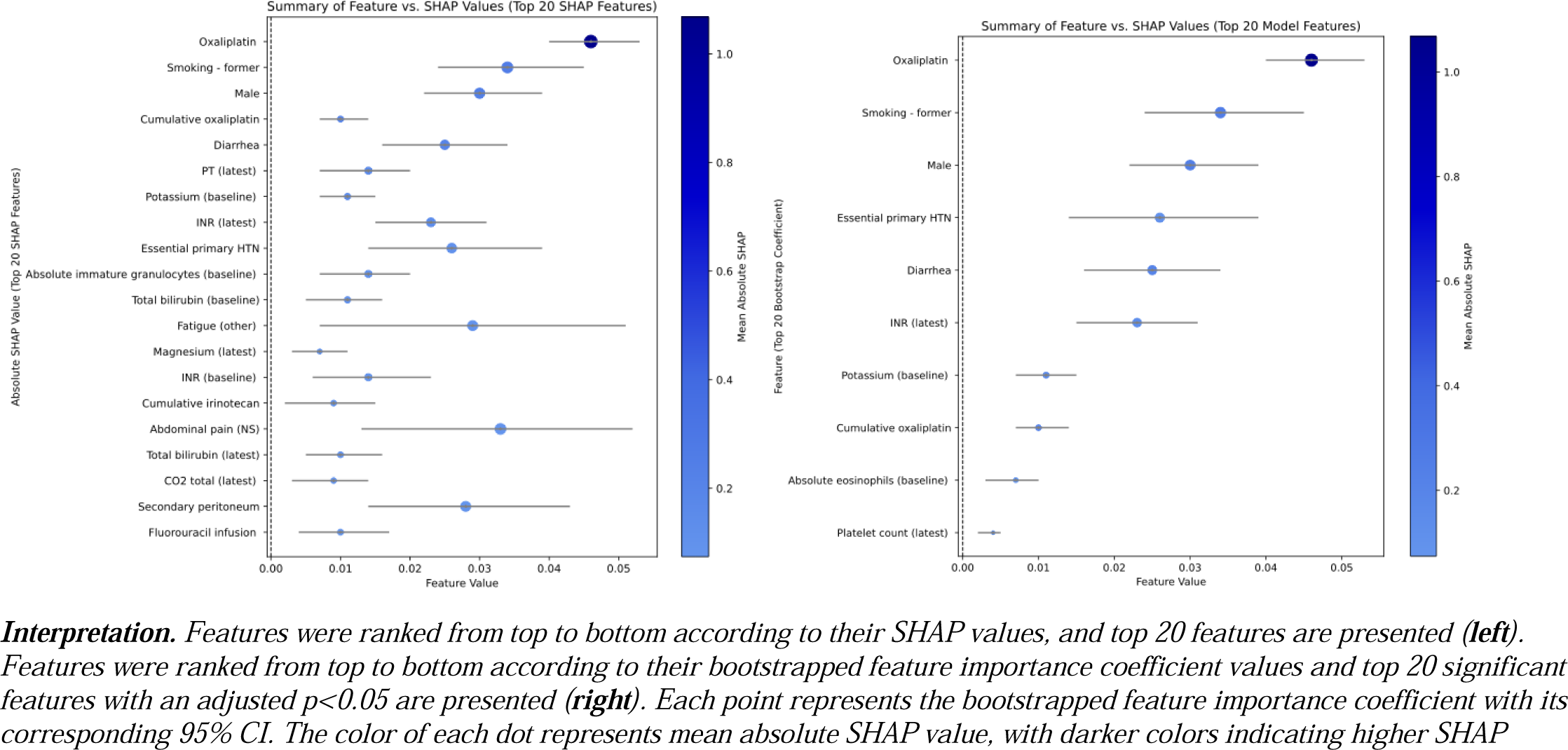
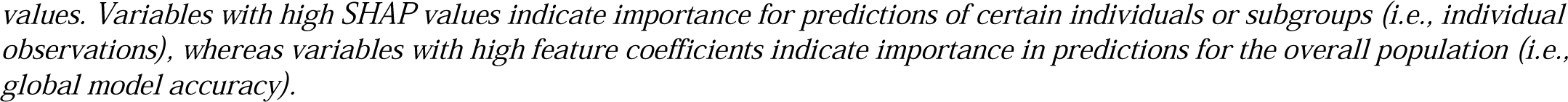
*FOLFIRINOX* Dose Modification/Reduction Feature Importance vs. SHAP Importance for Oxaliplatin.

## Discussion

Combination chemotherapy with FOLFIRINOX remains a cornerstone of treatment for eligible patients with PDAC, but its clinical benefit is limited by substantial toxicity and frequent need for dose modification.^22^ These modifications are typically related to drug-associated adverse events (AEs), yet in the absence of standardized dose-modification guidelines, decisions may vary substantially across providers and institutions due to differences in clinical judgment, risk tolerance, and local institutional practice patterns. Prior studies have demonstrated lack of consensus across oncology decision-making, including inter-institutional practice variation of approximately 10% in more than 20% of management decisions for patients with breast, lung, and colorectal cancer, as well as significant inter-physician variation in prescribing decisions under clinically comparable scenarios.^25,26^ In this context, understanding the patient characteristics associated with dose modification is essential, as these factors may represent clinically meaningful predictors of toxicity, markers of treatment tolerability, or signals of potentially unwarranted variation in clinical decision-making. In this retrospective longitudinal study, we developed and evaluated an EMR-based machine-learning framework to model cycle-specific FOLFIRINOX dose modification decisions in real-world oncology practice. Unlike models predicting a biologic outcome such as survival or progression, this framework models a treatment decision phenotype, which reflects both measured toxicity signals and clinician judgment.

In our study, over 60% of patients required at least one dose modification during the study period, and patients with dose modifications were significantly more likely to experience adverse outcomes during the study period, including higher rates of dehydration, polyneuropathy, and diarrhea. By comparison, previous studies conducted between 2013 and 2014 reported the rates of discontinuation of FOLFIRINOX regimens due to dose-limiting toxicities ranging from 29% - 42%.^27,28^ Although differences in study design, patient populations, and outcome definitions limit direct comparison, our findings from a contemporary tertiary-care cohort suggest that toxicity-related treatment modification remains highly prevalent in real-world practice and may represent a persistent, if not increasing, burden despite a decade of clinical experience with FOLFIRINOX. These findings underscore the need to better characterize real-world dosing patterns and evaluate how data-driven models can support earlier identification of patients at risk for treatment-limiting toxicity.

In FOLFIRINOX regimens, treatment with 5-fluorouracil forms the backbone of combination chemotherapy and was associated with lower treatment modifications compared to oxaliplatin and irinotecan across the cycles evaluated. The lower frequency of 5-fluorouracil modifications likely contributed to limited model performance, making its predictors difficult to interpret. By contrast, models for irinotecan and oxaliplatin identified clinically plausible predictors of dose modification, including markers of hepatic and renal function, coagulation measures, electrolyte abnormalities, clinical comorbidities, cumulative drug exposure, and demographic or behavioral characteristics such as sex and smoking status. Several findings support the face validity of the modeling framework. Total bilirubin was consistently associated with irinotecan dose modification, consistent with established concerns regarding irinotecan metabolism and hyperbilirubinemia. Similarly, cumulative and recent oxaliplatin exposure were strongly associated with oxaliplatin dose modification, reflecting known real-world patterns of cumulative toxicity and treatment de-escalation. The measured features do not entirely predict reductions, however, suggesting that there may be unmeasured effect modifiers of dose modifications (e.g., quality-of-life consideration) that mediate the relationship between known predictors and expected outcomes. These unmeasured factors could be at the level of providers (e.g., personal risk threshold, pre-existing attitudes, training background)^29,30^, institutions (e.g. department policies, ordering workflows, and local practice norms), patients (e.g., treatment preference and values, subtle declines in functional status)^31,32^, or the corresponding clinical context (e.g. scheduling uncertainties)^33^. Because many of these determinants are inconsistently documented or absent from structured EMR data, they may place an inherent ceiling on the achievable performance of purely data-driven models.

Several features were consistently associated with dose modifications across FOLFIRINOX components, offering insight into the real-world applicability of this approach. Female sex was associated with dose modification, which may reflect differences in treatment tolerance, optimal therapeutic dose, symptom reporting, clinician perception of risk, or patient preferences regarding quality of life. Prior work has shown better disease control and survival among female patients despite more frequent toxicity-related dose reductions, although the mechanisms underlying this association remain incompletely understood.^34,35^ We also identified an association between essential primary hypertension and oxaliplatin dose modification, which may reflect underlying comorbidity burden, vascular risk, or clinical frailty, though this finding requires validation in external cohorts. Importantly, several identified predictors, including liver enzymes, bilirubin, creatinine, and other routine laboratory measures, are readily available and inexpensive to monitor. These features may help identify patients at risk for treatment-limiting toxicity before they experience high-grade adverse events or require treatment discontinuation, after which they may no longer be able to benefit from chemotherapy.^36^ Despite the challenges of modeling complex treatment decisions, these consistent and clinically actionable associations highlight specific domains in which ML-based approaches may support individualized dosing and toxicity surveillance. Importantly, these models are best positioned to support risk stratification and earlier toxicity surveillance rather than replace clinician judgment.

This study represents a first step toward modeling the complex clinical decisions underlying real-world FOLFIRINOX dose modification, but it has several limitations. First, given the high-dimensional nature of the EMR-derived dataset, many predictors are likely correlated, introducing potential collinearity. Second, the absence of an external validation cohort limits the generalizability and transportability of our findings across health systems. Third, although our dataset included detailed longitudinal treatment data, dose modification may reflect toxicity, planned de-escalation, disease progression, patient preference, or clinician judgment, which may not be distinguishable in structured data. Finally, key drivers of dose modification, including patient-clinician communication, performance status, patient-reported symptoms, treatment goals, sociocultural factors, and quality-of-life considerations, are not routinely captured or are inconsistently documented in the EMR. This limits the ability of ML algorithms to learn reproducible patterns from structured clinical data alone and may explain the variable model performance observed in this study.

In conclusion, this study demonstrates both the promise and limitations of using longitudinal EMR-derived data to model real-world FOLFIRINOX dose modification decisions in PDAC. The framework identified clinically plausible and routinely available predictors of irinotecan and oxaliplatin modification, supporting the potential value of data-driven approaches for characterizing treatment de-escalation patterns and identifying patients at risk for treatment-limiting toxicity. However, variable model performance suggests that chemotherapy dosing decisions are only partially captured by structured clinical data and are likely mediated by patient-reported symptoms, functional status, quality-of-life considerations, clinician judgment, and local practice norms. Future oncology informatics efforts should prioritize structured documentation of dose-modification rationale, integration of patient-reported and functional outcomes, and validation across diverse practice settings to support interpretable, clinically useful decision-support tools.

## Data Availability

The data that support the findings of this study are derived from electronic health records and are not publicly available due to patient privacy and institutional restrictions. Access may be considered upon reasonable request and with appropriate institutional approvals.

## ACKNOWLEDGEMENTS

We dedicate this work to the memory of Dr. Atul Butte, whose contributions and mentorship were instrumental to this project.

