## Supplementary Appendix for "Real-World Dose Modifications for FOLFIRINOX in Pancreatic Cancer: Evaluating the Feasibility of A Machine-Learning Framework"

**Supplemental Table 1. Comprehensive List of Predictors Used in Models (Dose Modification and Clinical Outcomes)**

| **Features** | | | | |
| --- | --- | --- | --- | --- |
| Abdominal pain (NS) | Anxiety (NS) | Diarrhea | Marital status - SO | Race - Other, Unknown |
| Abnormal blood chemistry | Any neutropenia (baseline) | eGFR (baseline) | Marital status - unknown | Race - White |
| Abnormal EKG | Any neutropenia (latest) | eGFR (latest) | Max cycle | Secondary neoplasm - liver, bile-duct |
| Abnormal weight loss | Any polyneuropathy | Elevated WBC | MCH (baseline) | Secondary neoplasm (NS) |
| Absolute basophils (baseline) | Ascites | Essential primary HTN | MCH (latest) | Secondary peritoneum |
| Absolute basophils (latest) | AST (baseline) | Fatigue (other) | MCHC (baseline) | Severe neutropenia (baseline) |
| Absolute eosinophils (baseline) | AST (latest) | Fever (NS) | MCHC (latest) | Severe neutropenia (latest) |
| Absolute eosinophils (latest) | Bile duct obstruction | Fluorouracil infusion (last dose, cumulative dose, dose modification and/or reduction) | MCV (baseline) | Smoking - any |
| Absolute immature granulocytes (baseline) | BUN (baseline) | GERD | MCV (latest) | Smoking - daily |
| Absolute immature granulocytes (latest) | BUN (latest) | GERD w/o esophagitis | Mild neutropenia (baseline) | Smoking - former |
| Absolute lymphocytes (baseline) | Cancer antigen 19-9 (baseline) | Glucose - non-fasting (baseline) | Mild neutropenia (latest) | Smoking - never |
| Absolute lymphocytes (latest) | Cancer antigen 19-9 (latest) | Glucose - non-fasting (latest) | Moderate neutropenia (baseline) | Smoking - some |
| Absolute monocytes (baseline) | Cancer related pain | Hematocrit (baseline) | Moderate neutropenia (latest) | Sodium (baseline) |
| Absolute monocytes (latest) | Chloride (baseline) | Hematocrit (latest) | Nausea | Sodium (latest) |
| Absolute neutrophils (baseline) | Chloride (latest) | Hemoglobin (baseline) | Neoplasm-related pain | Thrombocytopenia (NS) |
| Absolute neutrophils (latest) | CO2 total (baseline) | Hemoglobin (latest) | Oxaliplatin (last dose, cumulative dose, dose modification and/or reduction) | Total bilirubin (baseline) |
| Acute postprocedural pain | CO2 total (latest) | Hispanic/Latino – no, yes, unknown | Pain (NS) | Total bilirubin (latest) |
| AE of antineoplastic immunosuppressive therapy | Constipation | Hypokalemia | Platelet count (baseline) | Total calcium (baseline) |
| Albumin (baseline) | Creatinine (baseline) | INR (baseline) | Platelet count (latest) | Total calcium (latest) |
| Albumin (latest) | Creatinine (latest) | INR (latest) | Potassium (baseline) | Vomiting |
| ALP (baseline) | Cumulative fluorouracil | Irinotecan (cumulative dose, last dose, dose modification and/or reduction) | Potassium (latest) | WBC count (baseline) |
| ALP (latest) | Cumulative irinotecan | Magnesium (baseline) | Protein - total (baseline) | WBC count (latest) |
| ALT (baseline) | Cumulative oxaliplatin | Magnesium (latest) | Protein - total (latest) |  |
| ALT (latest) | Current age | Male | PT (baseline) |  |
| Anion gap (baseline) | Cycle number | Marital status - divorced, widowed | PT (latest) |  |
| Anion gap (latest) | Day | Marital status - married | Race - Asian |  |
| Anorexia | Dehydration | Marital status - single | Race - Black/AA |  |

**Abbreviations:** AA: African-American, AE: adverse effect, ALP: alkaline phosphatase, AST: aspartate transaminase, BUN: blood urea nitrogen, CO2: carbon-dioxide, eGFR: estimated glomerular filtration rate, EKG: electrocardiogram, HTN: hypertension, GERD: gastroesophageal reflux disease, INR: international normalized ratio, MCV: mean corpuscular volume, MCHC: mean corpuscular hemoglobin concentration, NS: not specified, PT: prothrombin time, SO: significant other, WBC: white blood cell

**Note**: The above table includes the exhaustive list of features used across all of the models tested in the current analysis. However, the feature list for individual models varied based on outcome assessed and excluded variables that were highly correlated with the outcome (e.g., baseline dehydration in the model predicting future dehydration).

**Supplemental Table 2. Model Parameters**

| Model | Parameters |
| --- | --- |
| Logistic regression | - L2 regularization - Grid search regularization parameter C (0.01, 0.1, 1, 10, 100) |
| Random forest | - Number of trees: (50, 100) - Maximum depth: (10, 20, none) - Minimum samples for splitting: (5, 10) - Minimum samples at each leaf node: (2, 4) |
| XGBoost | - Number of boosting rounds: (50, 100) - Learning rate: (0.1) - Maximum depth: (3) - Minimum child weight: (1) |

**Abbreviations:** XGBoost: eXtreme gradient boosting

**Supplemental Table 3. Comparison of Characteristics for Patients with 0 or 1+ Dose Modifications or Reductions**

|  | Dose modifications across cycles  (Zero) | Dose modifications across cycles (1+) | *p-value* |
| --- | --- | --- | --- |
|  | 186 | 328 |  |
| **Baseline demographic characteristics** |  |  |  |
| Age at start of study | 59.70 (10.87) | 58.21 (12.42) | 0.173 |
| Male | 104 (55.9) | 170 (51.8) | 0.424 |
| Race (Asian) | 26 (14.0) | 78 (23.8) | 0.011 |
| Race (Black or AA) | 15 (8.1) | 7 (2.1) | 0.003 |
| Race (other/unknown) | 30 (16.1) | 52 (15.9) | 1 |
| Hispanic or Latino (yes) | 10 (5.4) | 31 (9.5) | 0.142 |
| Hispanic or Latino (unknown) | 12 (6.5) | 2 (0.6) | <0.001 |
| Marital status (married/significant other) | 128 (68.8) | 234 (71.3) | 0.616 |
| Marital status (unknown) | 6 (3.2) | 3 (0.9) | 0.116 |
| Smoking (any) | 11 (5.9) | 12 (3.7) | 0.334 |
| Smoking (former) | 72 (38.7) | 116 (35.4) | 0.509 |
| **Dosage information** |  |  |  |
| Cumulative 5-fluorouracil dose | 2212.37 (415.03) | 2450.46 (397.60) | <0.001 |
| Cumulative irinotecan dose | 145.91 (44.74) | 151.09 (43.35) | 0.199 |
| Cumulative oxaliplatin dose | 72.69 (19.90) | 72.31 (27.11) | 0.867 |
| **Baseline clinical characteristics** |  |  |  |
| Anemia | 61 (32.8) | 142 (43.3) | 0.025 |
| Diarrhea | 51 (27.4) | 150 (45.7) | <0.001 |
| Any polyneuropathy | 18 (9.7) | 114 (34.8) | <0.001 |
| Adverse effect of antineoplastic immunosuppressive therapy | 22 (11.8) | 111 (33.8) | <0.001 |
| Ascites | 20 (10.8) | 49 (14.9) | 0.229 |
| Thrombocytopenia | 14 (7.5) | 51 (15.5) | 0.013 |
| Nausea | 122 (65.6) | 243 (74.1) | 0.053 |
| Secondary neoplasm of liver or bile duct | 59 (31.7) | 154 (47.0) | 0.001 |
| Fatigue | 35 (18.8) | 119 (36.3) | <0.001 |
| GERD | 14 (7.5) | 29 (8.8) | 0.725 |
| Hypokalemia | 29 (15.6) | 72 (22.0) | 0.103 |
| Essential primary hypertension | 50 (26.9) | 96 (29.3) | 0.635 |
| Acute postprocedural pain | 19 (10.2) | 61 (18.6) | 0.017 |
| Elevated WBC | 16 (8.6) | 47 (14.3) | 0.078 |
| Dehydration | 134 (72.0) | 278 (84.8) | 0.001 |
| Neoplasm related pain | 47 (25.3) | 111 (33.8) | 0.054 |
| **Baseline laboratory data** |  |  |  |
| Complete blood count (CBC) |  |  |  |
| WBC count (x10E9/L) | 7.49 (3.32) | 7.84 (5.12) | 0.407 |
| Absolute monocytes (x10E9/L) | 20.99 (113.08) | 18.30 (117.23) | 0.8 |
| Absolute neutrophils (x10E9/L) | 138.07 (736.04) | 139.04 (883.25) | 0.99 |
| Absolute eosinophils (x10E9/L) | 6.38 (35.14) | 5.70 (41.91) | 0.852 |
| Absolute basophils (x10E9/L) | 1.27 (6.97) | 1.47 (9.80) | 0.807 |
| Absolute lymphocytes (x10E9/L) | 76.29 (351.17) | 68.64 (351.59) | 0.813 |
| Absolute immature granulocytes (x10E9/L) | 0.11 (0.42) | 0.09 (0.36) | 0.548 |
| Platelet count (x10E9/L) | 254.71 (109.28) | 263.95 (97.93) | 0.325 |
| Hemoglobin (g/dL) | 12.60 (1.75) | 12.79 (1.89) | 0.248 |
| MCV | 90.63 (5.72) | 89.21 (6.18) | 0.01 |
| Coagulation studies |  |  |  |
| PT | 13.41 (1.16) | 13.39 (1.44) | 0.875 |
| INR | 1.07 (0.10) | 1.07 (0.13) | 0.895 |
| Comprehensive metabolic panel (CMP) |  |  |  |
| Sodium (mmol/L) | 137.19 (3.24) | 137.31 (2.79) | 0.667 |
| Potassium (mmol/L) | 4.13 (0.47) | 4.02 (0.45) | 0.01 |
| Chloride (mmol/L) | 102.48 (3.62) | 102.51 (3.23) | 0.942 |
| CO2 (total) (mmol/L) | 25.64 (2.34) | 25.35 (2.56) | 0.205 |
| Glucose (non-fasting) (mg/dL) | 135.43 (52.07) | 128.88 (45.80) | 0.139 |
| Calcium (total) (mg/dL) | 9.09 (0.51) | 9.09 (0.57) | 0.955 |
| Protein (total) (g/dL) | 6.91 (0.59) | 6.96 (0.59) | 0.357 |
| Albumin (g/dL) | 3.63 (0.49) | 3.67 (0.47) | 0.403 |
| Liver panel |  |  |  |
| ALT (U/L) | 59.59 (87.14) | 61.19 (110.44) | 0.865 |
| AST | 45.72 (50.27) | 45.92 (60.72) | 0.969 |
| ALP (U/L) | 158.60 (166.27) | 151.33 (181.03) | 0.653 |
| Bilirubin (total) (mg/dL) | 1.52 (3.01) | 1.45 (2.78) | 0.771 |
| Special tests |  |  |  |
| Anion gap (%) | 9.14 (2.15) | 9.29 (2.27) | 0.457 |
| Magnesium (mg/dL) | 1.97 (0.16) | 1.99 (0.20) | 0.192 |
| Cancer antigen 19-9 (U/mL) | 4485.77 (9552.19) | 6877.73 (14106.29) | 0.04 |
| Kidney function |  |  |  |
| eGFR | 89.51 (19.51) | 93.14 (19.66) | 0.044 |
| Creatinine (total) (mg/dL) | 0.80 (0.21) | 0.78 (0.21) | 0.429 |
| BUN (mg/dL) | 13.52 (4.99) | 13.09 (4.99) | 0.354 |
| **Outcomes** |  |  |  |
| Dehydration | 0 (0.0) | 120 (36.6) | <0.001 |
| Nausea | 103 (55.4) | 211 (64.3) | 0.057 |
| Vomiting | 63 (33.9) | 143 (43.6) | 0.039 |
| Nausea or Vomiting | 104 (55.9) | 212 (64.6) | 0.063 |
| Any neutropenia | 38 (20.4) | 88 (26.8) | 0.13 |
| Polyneuropathy | 13 (7.0) | 105 (32.0) | <0.001 |
| Diarrhea | 40 (21.5) | 130 (39.6) | <0.001 |
| Death | 115 (61.8) | 188 (57.3) | 0.365 |
| Age at death | 61.42 (10.19) | 60.33 (12.50) | 0.451 |

**Abbreviations:** ALT: alanine transaminase, AST: aspartate transaminase, ALP: alkaline phosphatase, BUN: blood urea nitrogen, CO2: carbon-dioxide, eGFR: estimated glomerular filtration rate, GERD: gastroesophageal reflux disease, INR: international normalized ratio, MCH: mean corpuscular hemoglobin, MCV: mean corpuscular volume, PT: prothrombin time, RBC: red blood cell, WBC: white blood cell
